# Epigenetic Responses to Abusive versus Accidental Injuries in Children: A Cross-sectional Epigenome Wide Association Meta-analysis

**DOI:** 10.64898/2026.02.02.26345419

**Authors:** Kyle A. Campbell, Audrey Raut, Kelsey Julian, Kim Kaczor, Kathi Makoroff, Todd M. Everson, Mary Clyde Pierce

## Abstract

Child maltreatment is a leading cause of pediatric morbidity and mortality, potentially propagated by DNA methylation (DNAm) changes. We conducted an EWAS meta-analysis (n=175, 554,979 Illumina EPICv1/EPICv2 sites) in buccal swabs from three hospital-based studies of children with traumatic injuries, stratified by study group to include 1) any traumatic injury, 2) fractures, and 3) traumatic brain injuries. Empirical bayes-moderated linear models tested differential DNAm with M-values, followed by near-promoter gene set enrichment analysis. Abuse was associated with methylation at 11 sites (q<0.10), including enhancers of neuroblast differentiation-associated *AHNAK*, immunomodulators *SCGB1A1* and *CCL26,* exon 5 of *LAMP1*, essential for lysosomal function and cytotoxicity, and *RGS7*, a GTPase essential for synaptic transmission. Enriched biological processes included cranial skeletal system and connective tissue development, neural structure and function, immune regulation, gene expression, and metabolism. Our findings suggest that early abuse may epigenetically affect both proximal injury responses and longer-lived systemic biological dysregulation.

## INTRODUCTION

Child abuse is a leading cause of pediatric morbidity and mortality, affecting more than 550,000 children in the United States each year.^1^ The profound harms of maltreatment extend beyond the immediate manifestations of the abusive injuries themselves, increasing a child’s risk for negative physical and mental health outcomes across the life course.^2–5^ However, the mechanisms linking child abuse to latent deleterious health issues such as cancer, diabetes, cardiovascular disease, autoimmune conditions, psychiatric disorders, and even premature death, have yet to be elucidated.

A growing body of evidence implicates the epigenetic process of DNA methylation (DNAm) as a key driver in the biological embedding of early traumatic experiences.^6^ DNAm regulates gene expression throughout the body in response to physical, behavioral, and socioemotional stimuli in the environment; these biological responses can be adaptive or maladaptive. Multiple studies have found child maltreatment to be associated with altered patterns of DNAm that may subsequently serve as mechanisms of disease.^7^ Much of this work has focused on candidate genes and pathways, especially those involved in stress response regulation (i.e.. HPA axis genes), and demonstrate an important role for epigenetic responses to trauma.^5,8–11^ Epigenome-wide association studies (EWAS) of childhood maltreatment have also identified differentially methylated loci outside of canonical stress-response pathways.^7^ However, the majority of EWAS studies 1) were performed using retrospective ascertainment of childhood maltreatment, often years after its occurrence, 2) included various forms of maltreatment, and 3) were individual studies rather than meta-analyses.^7^ Compelling questions therefore remain as to whether, when, and how early experiences of physical abuse induce epigenetic alterations that affect development and function through interactive pathways, shaping trajectories of health and disease.

Given that that early maltreatment has been linked to a broad range of long-term health outcomes,^2–5^ further epigenome-wide analyses are needed to capture the full scope of biological embedding. We therefore conducted an epigenome-wide association study (EWAS) meta-analysis incorporating DNAm data from three studies of young children at the time of presentation with traumatic injuries. The objectives of this study were to 1) identify differences in DNAm between those with abusive versus accidental injuries, and 2) examine which biological pathways may be affected by differential methylation between the two groups.

## METHODS

### Study Design and Composition

We conducted three independent observational studies exploring DNA methylation changes among young children with traumatic injuries. The first was a cross-sectional study of children 0-3.99 years of age who presented to the pediatric Emergency Department (pED) at Ann & Robert H. Lurie Children’s Hospital of Chicago or Cincinnati Children’s Hospital between August 2015 and October 2018 and were diagnosed with a traumatic injury. The second was a cross-sectional study of children 0-11.99 months of age who presented to Ann & Robert H. Lurie Children’s Hospital pED between October 2020 and July 2023 and were diagnosed with a long-bone fracture. The third was a cohort study of children 0-11.99 months of age who presented to Ann & Robert H. Lurie Children’s Hospital between October 2021 and June 2022 and were diagnosed with a traumatic brain injury (TBI).

For all three studies, trained staff collected standardized data including: demographics; history and physical exam findings; imaging and consultation findings; injury type, severity, and outcome; past medical history; and psychosocial risk factors (PRFs) present in the child’s caregiving environment, including substance abuse, domestic violence / intimate partner violence, untreated mental health problems, and prior involvement of law enforcement or child protective services.^12^ PRF scores ranging from 0-5 were calculated by summing the presence (1) or absence (0) of 5 psychosocial risk factors. Trained study staff also collected buccal swabs for DNA methylation measures at the time of presentation to the hospital. Each case was classified clinically as abuse, accident, or indeterminate by a child abuse assessment team (TBI study) or medical expert panel (trauma and fracture studies).^13^

All studies were reviewed and approved by the Institutional Review Boards at Lurie Children’s Hospital and/or Cincinnati Children’s Hospital. For patients undergoing abuse evaluations, waivers of authorization were granted to allow the research team to abstract data while avoiding interference with the evaluation process. For patients not undergoing an abuse evaluation, the research team obtained written informed consent for data collection, with fields designed to parallel the more extensive and detailed dataset collected as standard of care for abuse consultations. Verbal consent for buccal swab sampling was obtained from all parents prior to specimen collection. All data and epigenetic samples were deidentified.

### DNA methylation measurement, preprocessing, and quality control

Buccal cells were collected via SK-1S Isohelix swabs with Isohelix Dri-Capsules (Boca Scientific) for DNAm measurements. DNA was extracted and quantified with the Qubit Fluorometer (Thermo Fisher, Waltham, MA) and then aliquoted into standardized concentrations to yield 500ng. Samples were plated randomly across 96-well plates across study batches. For the traumatic injuries study, Northwestern University’s (NU) Genomics (NUSeq) Core Facility performed bisulfite modification using the EZ DNA Methylation Kit (Zymo Research, Irvine, CA), and measured DNAm throughout the genome with the Illumina MethylationEPIC Beadarray (Illumina, San Diego, CA) following the manufacturer’s protocol. For the TBI and Fracture studies, the Emory University Integrated Genomics Core performed bisulfite conversion using the EZ DNA Methylation Kit (Zymo Research, Irvine, CA) and measured DNAm throughout the genome using the Illumina MethylationEPIC Beadarray Version 2.0 (Illumina, San Diego, CA) following the manufacturer’s protocol.

We processed and performed quality control of DNA methylation data in R statistical software (version 4.3.3) for each study. Raw DNA methylation data EPICv1 (trauma study) and EPICv2 (TBI and fracture studies) idats were preprocessed and managed separately in minfi R package v1.48.0^14^ and ewastools v1.7.2^15^ with default Illumina hg38-based manifests. We preprocessed raw DNA methylation data with Noob background correction and dye-bias normalization. To assess quality control, we checked each sample for the 17 Illumina control metrics, poor detection p-values averaging > 0.05 across all loci, and sex or genotype mismatches. Samples that failed multiple quality control checkpoints were excluded. We further excluded each probe for >1% failure across samples for detection p-value > 0.01, cross-reactive or SNP-related probes^16^, and sex-specific probes. To eliminate genetic or other unwanted sources of DNA methylation signal in beta values, we removed non-outlier gap signals of 10% or greater present in 5% or more of samples (20% in the fracture subgroup to ensure comparability with larger samples) with gaphunter in minfi v1.48.0.^14^ Finally, to correct for type I vs. type II probe bias, we also performed Beta MIxture Quantile dilation normalization^17^ via the ChAMP package v2.22.0^18^. All downstream DNA methylation analyses used the M-value or the methylation level of a given site. DNA methylation quality control is summarized in **Supplementary Figure 1**. For all included samples, we describe the number and frequency of samples by study source, fetal sex, cell type, Illumina array type, and cell deconvolution method.

### Estimation of DNA methylation-based features

Unless otherwise noted, all procedures were stratified by study group (trauma, fracture, or TBI) and conducted with default software settings. To account for potential unmeasured age-specific epigenetic effects, cell type heterogeneity, and other sources of unmeasured confounding, we used DNA methylation data and covariate data to estimate buccal swab cell proportions with methylation rate values and surrogate variables with DNA methylation M-values. Because all external references used to estimate epigenetic features were trained on EPICv1 microarrays, for EPICv2 samples, we lifted over probes from EPICv2 to EPICv1 using the sesame R package v1.20.0.^19^ To calculate cell heterogeneity, we estimated epithelial vs. immune cell proportions using beta values in EpiDISH R package v2.18.0^20^ with the robust partial correlation algorithm using a previously published reference trained on saliva samples from children aged 7-16 years old.^21^ Due to systematic differences in cell composition, we excluded samples that were estimated to be less than 50% epithelial. To account for potential unmeasured confounding based on *a priori* knowledge, surrogate variables were created from M-values adjusted for sex (dichotomous), age (continuous, months), psychosocial risk score (sum 0-5 factors present), insurance status (private vs. none/public, dichotomous), estimated epithelial cell proportion (continuous), race (white vs. non-white), ethnicity (Hispanic vs. non-Hispanic, dichotomous), and batch (trauma study only, 1 vs. 2, dichotomous) using the sva R package v3.50.0 with 20 iterations.^22^ To ensure surrogate variables were only capturing unmodelled variation, we tested the association between surrogate variables and covariates or the exposure with analysis of variance or correlation tests. We also conducted surrogate variable-wise leave-one-out analyses to prune uninformative surrogate variables and optimize a genomic inflation factor closest to one.

### Statistical analysis

Primary linear epigenome-wide association analyses with empirical Bayes-moderated standard errors^23^ were stratified by study group: traumatic injuries among children <4 years of age (abuse=22, accident=39), TBI among children <1 year of age (abuse=20, accident=73), and fractures among children <1 year of age (abuse=7, accident=11), before meta-analysis. For harmonization, the EPICv1 trauma study was lifted over to EPICv2 loci with sesame R package v1.20.0.^19^ Each subtype epigenome-wide association study as well as their meta-analysis used M-values^24^ and were assessed for inflation with a quantile-quantile plot and genomic control inflation factor. Because we did not identify systemic evidence of effect heterogeneity in a random effects meta-analysis, we conducted a fixed effects meta-analysis. We independently filtered low variance probes^25^ defined by median absolute deviation less than 1% on the beta value scale across all three individual EWASs, yielding a primary analysis across 408,756 common EPICv2 loci. We performed rank-based gene set enrichment analysis of epigenome-wide association results to identify biological processes enriched among differential methylation results at loci annotated to or near promoters and of gene targets of loci located within cis-regulatory elements in a secondary analysis. Models were adjusted for sex, insurance, psychosocial risk score, cell composition, race, ethnicity, and surrogate variables. The fracture group analysis was only adjusted for sex, age, and surrogate variables because the fully adjusted model could not be fit due to a lack of support across additional covariates. Statistical significance was assessed at nominal p-value < 0.05 or false discovery rate adjusted^26^ q < 0.25 for genome-wide association tests unless noted otherwise; we highlight those results that yield q < 0.10 as our most robust findings.

### Annotation of meta-analysis results and gene set enrichment analysis

To establish gene-to-probe mapping, epigenome-wide association study results were annotated to the Illumina EPICv2 manifest v20a1.hg38. To identify biological gene ontologies overrepresented among abuse-associated loci annotated near promoters (classified as within 200-1500 base pairs of the transcription start site, or within exon one or the 5’ untranslated region), we performed gene set enrichment analysis^27^ against pathways ranging from 20 to 1000 genes after adjusting p-values with robust rank aggregation^28^ in methylGSA v1.20.0.^29^ If a locus was annotated to multiple genes, we selected the most upstream feature as the annotated gene. For additional annotation information we consulted GencodeV47^30^ in the UCSC Genome Browser^31^ (accessed January 2025) and the GeneHancer database (accessed October 20^th^, 2025) to annotate DNAm loci to cis-regulatory elements (CREs) and their target genes.^32^ The same analysis pipeline was used to test CRE gene targets for biological process enrichment. If multiples genes mapped to different DNAm loci, we selected the locus with the lowest p-value in the primary analysis.

## RESULTS

### Study Samples and Demographics

In total, 175 patients were included: 61 with traumatic injuries (abuse=22, accident=39), 18 with fractures (abuse=7, accident=11), and 93 with TBI (abuse=20, accident=73). There were no consistent (p < 0.05) bivariate associations between abuse determination and sex, estimated epithelial cell proportion of buccal samples, race, or ethnicity for any study group (**Table 1**). We observed consistent associations between abuse determination and PRF scores in the traumatic injury and TBI groups, and between abuse determination and insurance status in all study groups. Within each study group, median PRF scores were approximately two points higher among patients with abusive compared to accidental injuries, though this difference did not meet statistical significance in the fracture group. Patients with abusive injuries were more likely to have public insurance or no insurance compared to patients with accidental injuries, who were more likely to have private insurance (p < 0.05). Age and abuse determination were strongly associated in the fracture group, with a median age of 2.52 months among patients with abusive injuries and 9.48 months among those with accidental injuries (p = 0.007).

**Table 1.**
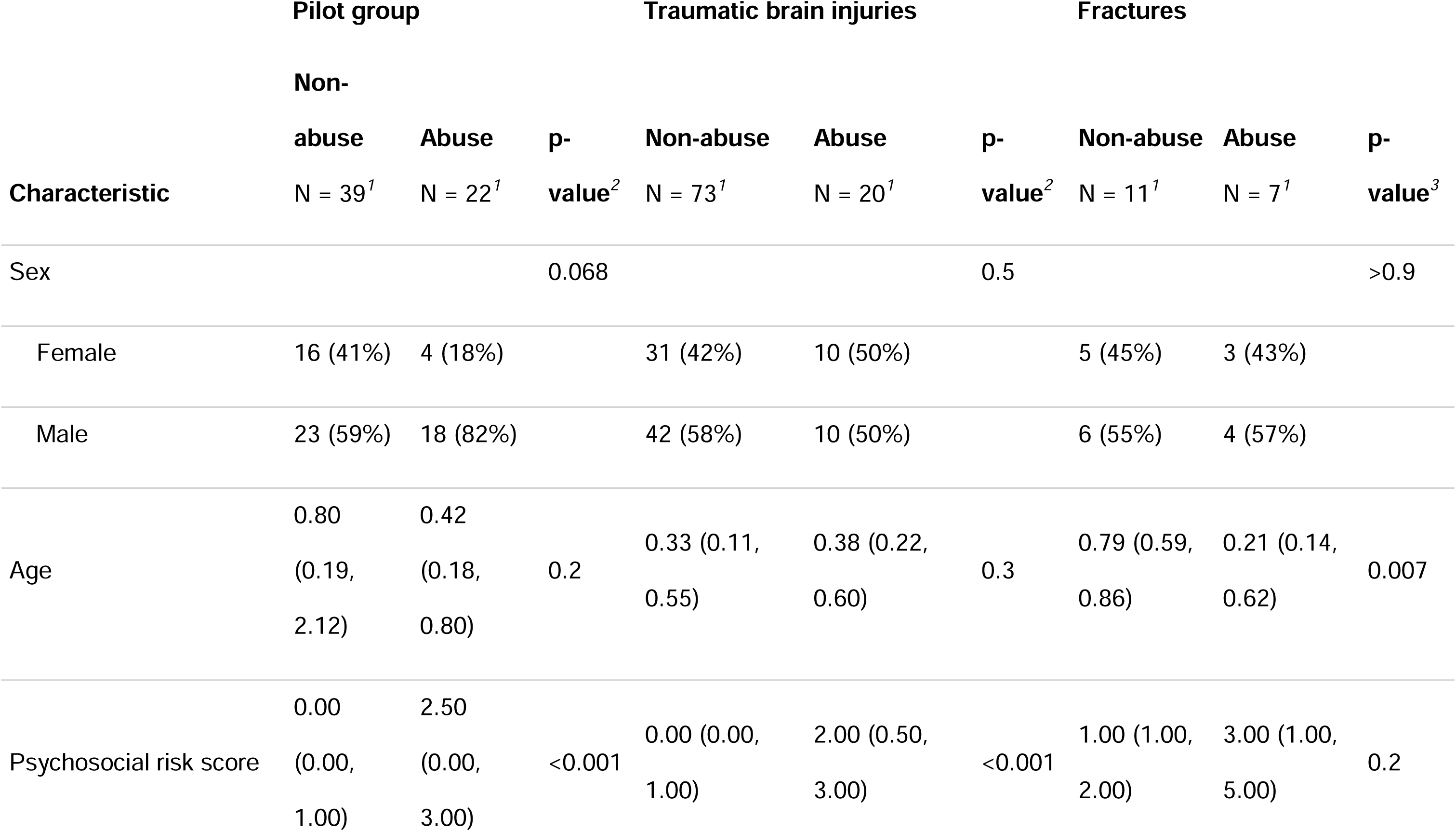
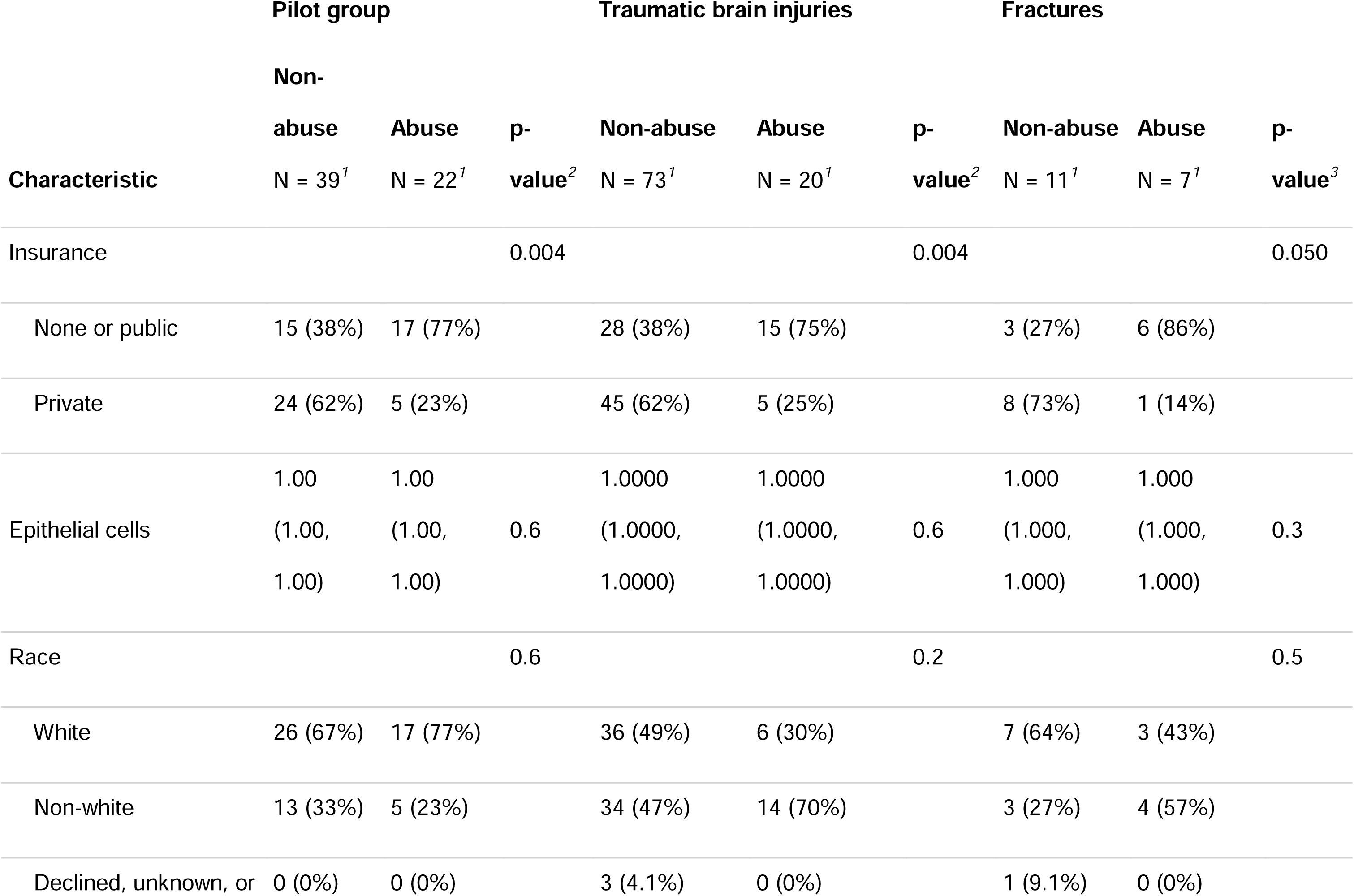
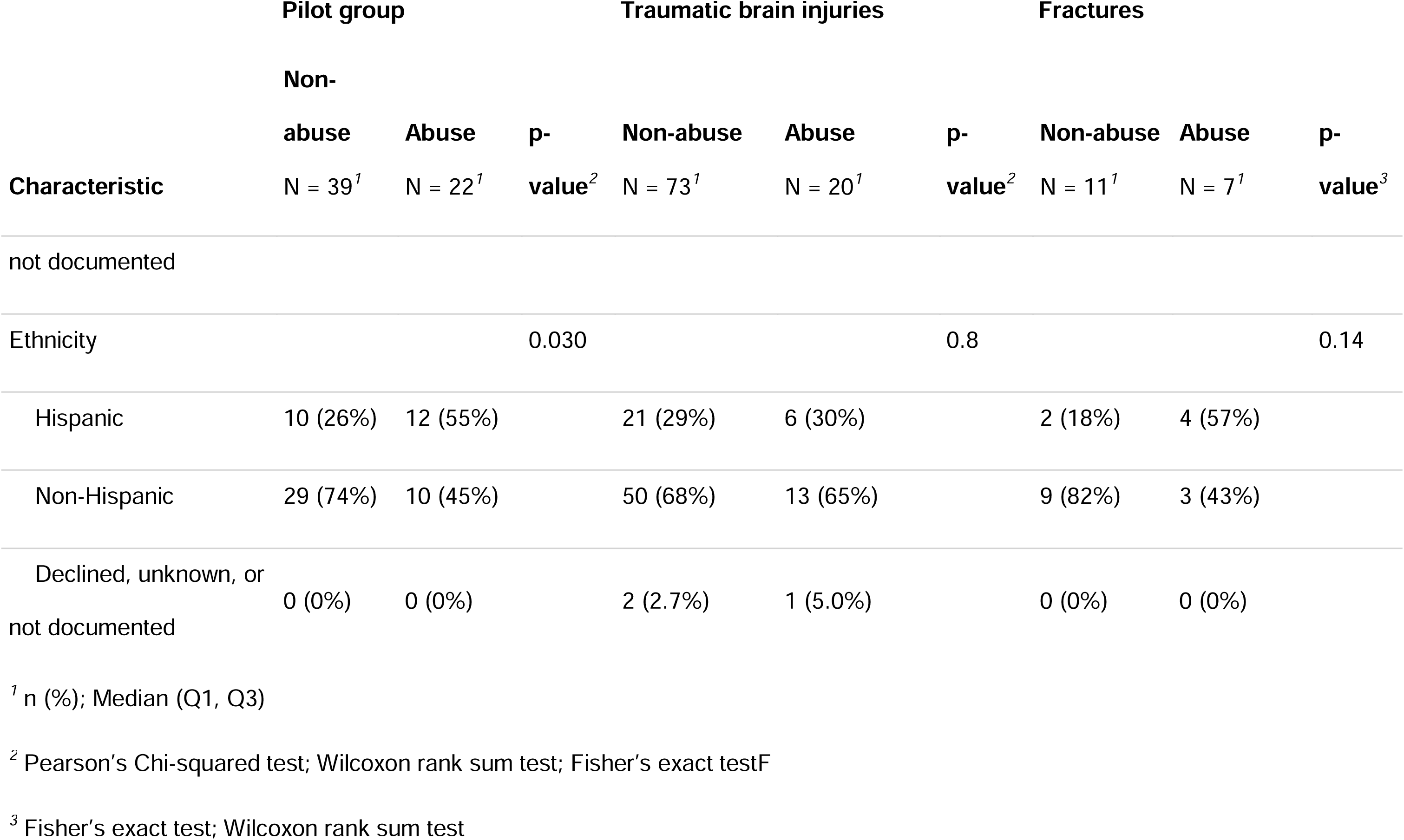
Demographic and covariate distributions stratified by subgroup and compared by exposure status.

To identify novel epigenetic DNA methylation changes associated with abuse in the context of different traumatic injuries, our initial epigenome-wide association analyses were stratified by study group: traumatic injuries, fractures, and TBI, as outlined below.

### Traumatic Injuries Group EWAS

In the traumatic injuries group, after DNA methylation quality control procedures (**Supplementary Figure 1, Trauma group**), our sample included 61 patients up to four years of age (abuse=22, accidental=39), with analysis across 578,406 DNA methylation loci as measured on the EPICv1 microarray and lifted over to the EPICv2 microarray manifest. We imputed ethnicity for 1 observation and psychosocial risk score for 2 individuals. We performed an epigenome-wide association study by regressing site-specific DNA methylation M-values on abuse determination, adjusted for sex, age, psychosocial risk score, insurance status, estimated epithelial cell proportion, race, ethnicity, batch and 10 surrogate variables to account for unmeasured confounding (**Supplementary Table 1, Trauma group**). With a genomic inflation factor of 0.94, our results indicate that systematic bias was minimal, suggesting that population structure, technical artifacts, and other potential confounders were well-controlled in our analysis (**Supplementary Figure 2A**). No loci achieved epigenome-wide significance after false discovery rate adjustment (q < 0.05); however, 44 loci were nominally associated with abusive injury at p < 0.0001 (**Supplementary Figure 3**). Methylation at the top locus, cg13150171_BC11, was associated with a log_2_ fold-change of –0.59 95% CI (–0.83, –0.36) among abusive compared to accidental injuries; the average methylation rate decreased from 6.7% to 4.6% and annotated to exon 1 and 3’ untranslated region (UTR) of Transcriptional Co-Activator 4, *CITED4*.

Despite the lack of a robust genome-wide signal, biological process gene set enrichment analysis of promoter-associated DNA methylation loci annotated to within 1500bp of a gene’s transcription start site, exon 1, or 5’ UTR revealed that the axonal growth cone pathway was overrepresented (q < 0.05), driven by over– and undermethylation at 15 loci annotated to *SCN11A*. An additional 11 pathways were enriched at q < 0.10, including endoplasmic reticulum stress, upregulation of Rho protein signal transduction, chromosome localization, oligodendrocyte differentiation, and phospholipid biosynthesis. A further 30 pathways were significantly enriched at q < **0.25 (**Supplementary Table 2, Trauma group).

### Fracture Group EWAS

In the fracture group, after DNA methylation quality control procedures (**Supplementary Figure 1, Fracture group**), our sample included 18 individuals under one year of age (abuse=7, accident=11), analyzed across 737,068 loci on the EPICv2 microarray. We imputed race for one observation. We performed an epigenome-wide association study by regressing site-specific DNA methylation M-values on abuse determination, adjusted for sex, age, and three surrogate variables to account for unmeasured confounding (**Supplementary Table 1, Fracture group**). Our model was moderately inflated at a genomic inflation factor of 1.12 (**Supplementary Figure 2C**), and 114 loci were nominally associated with abusive fractures (**Supplementary Figure 5**). Methylation at the top locus, cg07239227_TC21, was associated with a log_2_ fold-change of –2.17 95% CI (–2.75, –1.58) (q = 0.28) among abusive compared to accidental fractures; the average methylation rate decreased from 96.0% to 84.0% and annotated to exon 2 of *ZNF579*.

EWAS results for abusive versus accidental fractures were enriched for 68 biological processes at q < 0.05 across a variety of domains including pathways highly relevant to fracture injuries such as osteoblast differentiation, megakaryocyte development, regulation of hemopoiesis, epithelial morphogenesis, and bone development. Other enriched pathways were related to altered immune responses, epigenetic regulation such as chromatin binding and histone kinase activity, and one-carbon metabolic processes, peptide responses, hormone responses, germ cell development, carbohydrate metabolic processes, non-coding RNA metabolic processes, and ensheathment of neurons. A further 124 pathways were enriched at q < 0.10, including ossification (q = 0.05) and regulation of bone remodeling (q = 0.06) (**Supplementary Table 2, Fracture group**).

### TBI Group EWAS

In the TBI group, after DNA methylation quality control procedures (**Supplementary Figure 1, TBI group**) and exclusion of three accidental injuries missing psychosocial risk factor information, our sample included 93 individuals under one year of age (abuse=20, accident=73), analyzed across 737,068 loci on the EPICv2 microarray. We performed an epigenome-wide association study by regressing site-specific DNA methylation M-values on abuse determination, adjusted for sex, age, psychosocial risk score, insurance status, estimated epithelial cell proportion, race, ethnicity, and five surrogate variables to account for unmeasured confounding (**Supplementary Table 1, TBI group**). Our model was well-adjusted with a genomic inflation factor of 1.01 (**Supplementary Figure 2B**), and 70 loci were nominally associated with abuse determination at p < 0.0001 (**Supplementary Figure 4**) Methylation at the top locus cg02967813_TC21 was associated with a log_2_ fold-change of –0.79 95% CI (–1.06, –0.51) (q = 0.18) among children with abusive compared to accidental injuries; the average methylation rate decreased from 98.2% to 97.0% and annotated to intron 3 of *COLEC11*, a lectin involved in innate immunity, apoptosis, and neural crest migration.^33^

Enrichment analysis of differential methylation results in the TBI group identified 11 enriched biological pathways at q < 0.10 related to carboxylic acid transmembrane transporter activity, retrograde axonal transport, long-chain fatty-acyl-CoA metabolic process, cellular response to peptide hormone stimulus, G protein-coupled peptide receptor activity, and gamete generation. An additional 37 pathways were enriched at q < 0.25, including integrin activation, synapse organization, endochondral ossification, positive regulation of epidermis development, response to interleukin-4, and dendritic cell differentiation (**Supplementary Table 2, TBI group**).

### EWAS Meta-Analysis

To identify a robust cross-traumatic injury epigenomic DNA methylation signal for abusive versus accidental injuries, we conducted a meta-analysis across all three groups. Because heterogeneity statistics were well-controlled with only 429 of 555,135 sites showing effect heterogeneity (q < 0.05) in a random effects model, we present a fixed effects model. We independently filtered 146,379 low variance probes defined by median absolute deviation less than 1% on the beta value scale in all three individual EWASs, yielding a primary analysis across 408,756 common DNA methylation loci for 49 abusive and 123 accidental traumatic injuries (**Figure 1, Supplementary Table 3**). Inflation was well-controlled model at 0.99 before low variance filtering and was slightly inflated after filtering with a genomic inflation factor of 1.04 (**Supplementary Figure 2D**).

**Figure 1.**
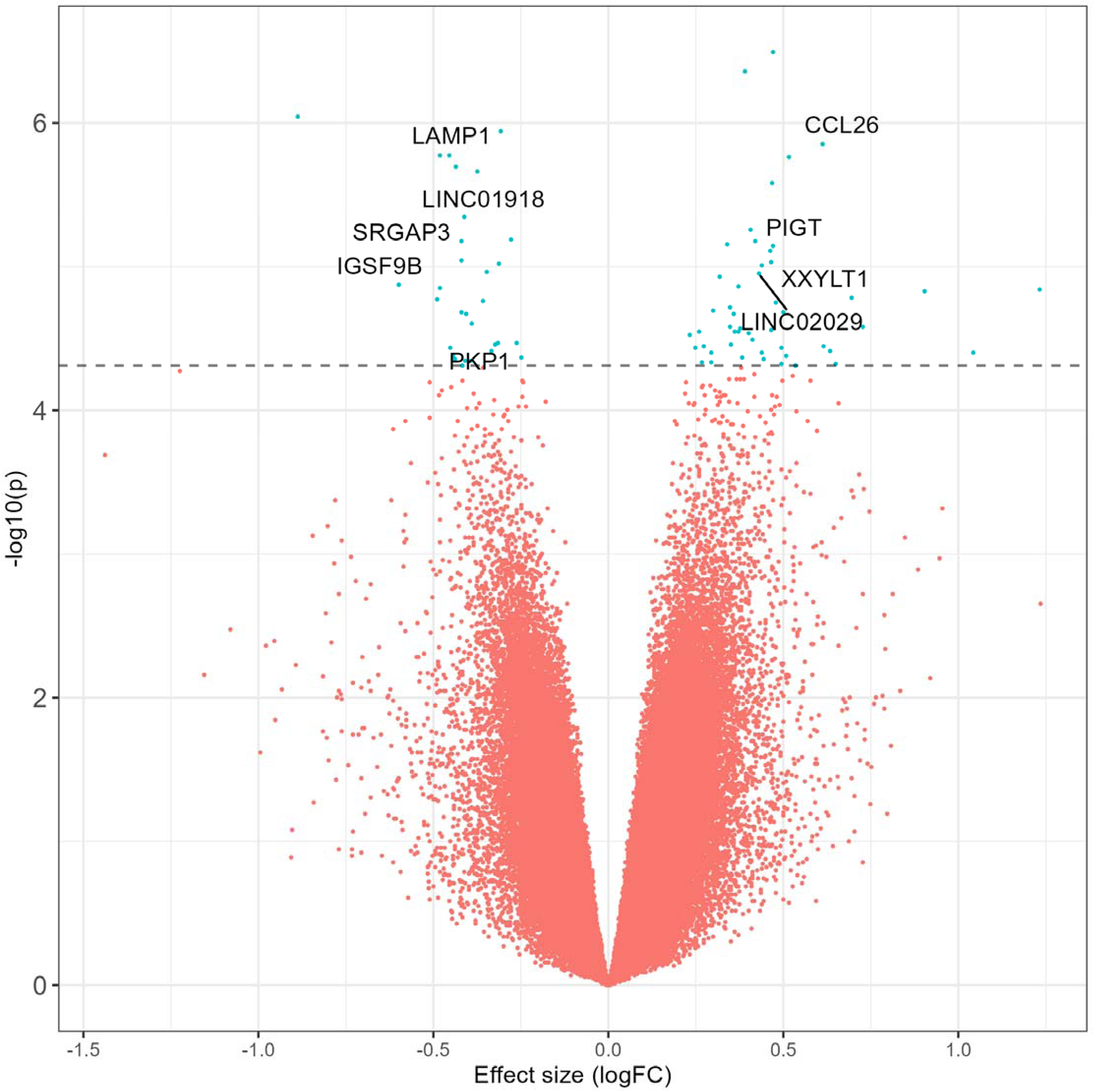
Volcano plot summarizing the differentially methylated loci in the epigenome-wide association study meta-analysis of abusive versus accidental traumatic injuries.

A total of 80 loci were differentially methylated at q < 0.25, and we identified 11 differentially methylated loci at q < 0.10 (**Table 2**). Effect magnitude and direction were generally consistent across individual EWASs and the meta-analytic estimate (**Figure 2**) among these 11 loci. Methylation at the top locus, cg24890736_BC21, was associated with a log2 fold-change of 0.47 95% CI (0.29, 0.65) (q = 0.09) compared to accidental injuries. The average methylation rate increased from 91.1% to 93.4% and annotated to an Alu short interspersed nuclear element repeat, potentially relevant for genomic stability. Methylation at the next hit, cg20544725_BC21, was higher with a log_2_ fold-change of 0.39 95% CI (0.24, 0.54) (q = 0.09) among abusive compared to accidental injuries. The average methylation rate increased from 87.0% to 90.0% and annotated to intron 1 of *ENSG00000295816*, an enhancer long non-coding RNA antisense to neuroblast differentiation-associated *AHNAK*. Other top differentially methylated loci mapped to *LAMP1*, a regulator of lysosomal function and autophagy,^34^ as well as natural killer cytotoxicity;^35^ *CCL26*, a chemokine that recruits eosinophils and other immune cells;^36^ and *RGS7*, a GTPase-activating protein that is highly expressed in the brain and essential for inhibiting neuronal G-protein-coupled receptors in synaptic transmission.^37^

**Figure 2.**
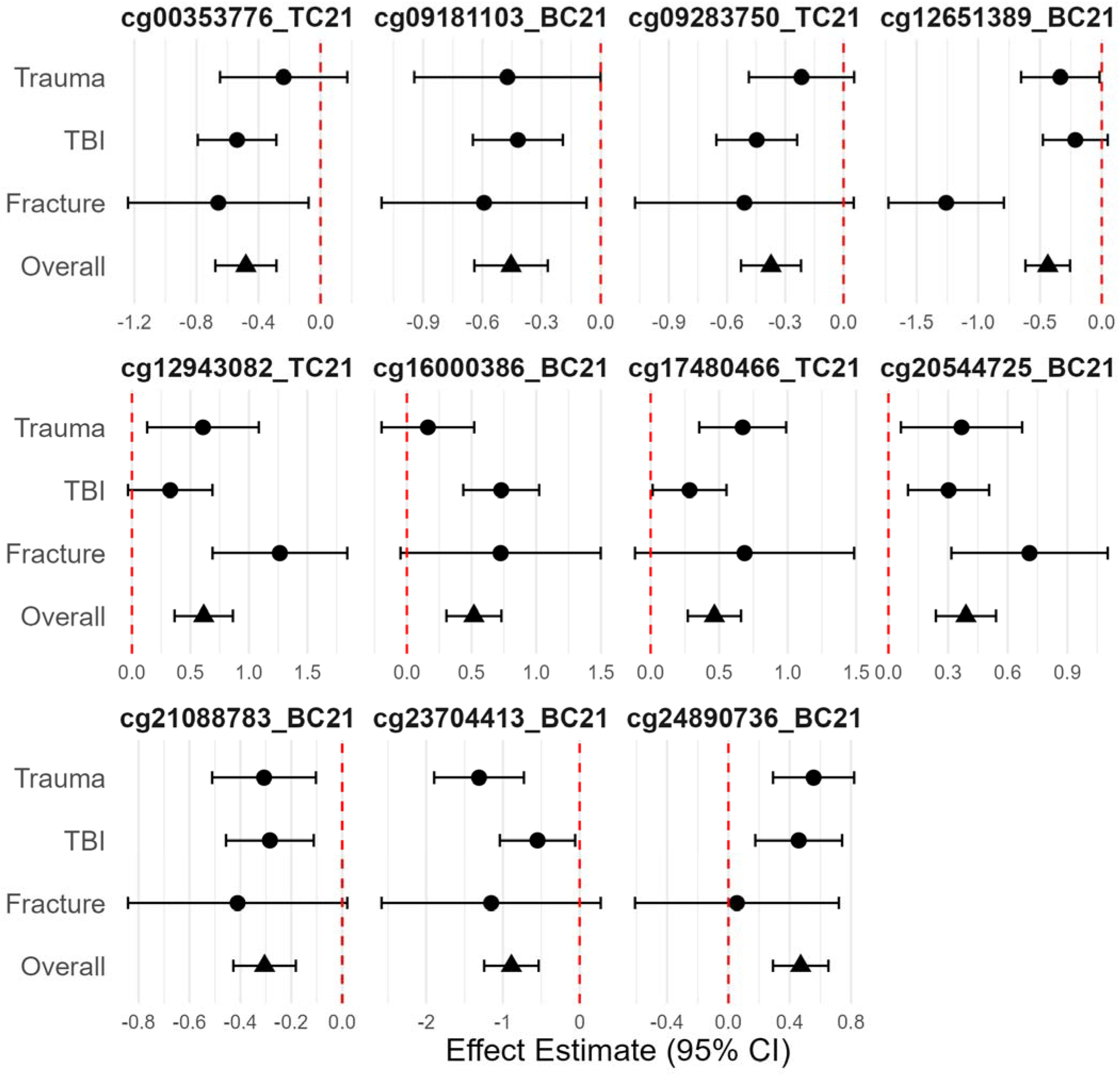
Forest plot summarizing group-specific effect estimate and the overall meta-analytic estimate for the 11 top differentially methylated loci in the epigenome-wide association study meta-analysis of abusive versus accidental traumatic injuries.

**Table 2.** Top 11 meta-analytic EWAS hits that meet false discovery rate-adjusted q-value of 0.10.

| Probe ID | p-value | Average Beta Value (%) | Effect size (Beta Value) | Biological Significance | Gene | Gene element |
| --- | --- | --- | --- | --- | --- | --- |
| cg24890736_BC21 | 3.21E-07 | 0.911 | 0.023 | Alu SINE, genomic stability | N/A | N/A |
| cg20544725_BC21 | 4.39E-07 | 0.870 | 0.028 | *AHNAK/SCGB1A1* enhancer, neurodifferentiation | ENSG00000295816 | Intron 1 |
| cg23704413_BC21 | 9.08E-07 | 0.737 | -0.135 | *PTPRN2* is a neuroendocrine gene | *PTPRN2* | Intron 6 |
| cg21088783_BC21 | 1.14E-06 | 0.384 | -0.049 | Predicted cis-regulatory element | ENSG00000298479 | Intron 1 |
| cg12943082_TC21 | 1.39E-06 | 0.863 | 0.043 | *CCL26* enhancer, immune cell recruiter | *CCL26* | TSS1500 |
| cg00353776_TC21 | 1.66E-06 | 0.945 | -0.020 | lncRNA antisense to *GZMK* | ENSG00000240535 | Intron 2 |
| cg09181103_BC21 | 1.71E-06 | 0.969 | -0.011 | lysosome, autophagy, natural killer cytotoxicity | *LAMP1* | Exon 5 |
| cg16000386_BC21 | 1.72E-06 | 0.585 | 0.084 | SINE near myocardial infarction ncRNA | *MIATNB* | Intron 11 |
| cg12651389_BC21 | 2.03E-06 | 0.960 | -0.013 | Unknown ncRNA | ENSG00000229771 | Intron 5 |
| cg09283750_TC21 | 2.18E-06 | 0.948 | -0.014 | *FAM135B*, previously associated with emotional abuse | *FAM135B* | TSS2500 |
| cg17480466_TC21 | 2.63E-06 | 0.980 | 0.006 | *RGS7*, neuron-expressed regulator of GPCRs | *RGS7* | Intron 10 |

Biological process gene set enrichment analysis of near-promoter DNA methylation loci identified 5 highly enriched pathways (q < 0. 05) (**Figure 3**), including cranial skeletal system development, response to biotic stimulus and lipopolysaccharide, and DNA-binding transcription activator activity. Among the 147 pathways enriched at q < 0.25 (**Supplementary Table 4**), several biologically relevant themes emerged that may relate to injury mechanisms, development, and dysregulation of critical body systems. Neurodevelopment and potential brain injury processes were prominent including forebrain neuron generation and synaptic structure, along with dopamine and monoamine signaling. Additional enrichment in chromatin remodeling and DNA repair, endocrine hormone and growth factor signaling, developmental pathways (e.g., mesoderm and connective tissue development), and immune, adhesion, and redox responses points to coordinated epigenetic regulation, immune system, physiological stress adaptation, and tissue repair mechanisms.

**Figure 3.**
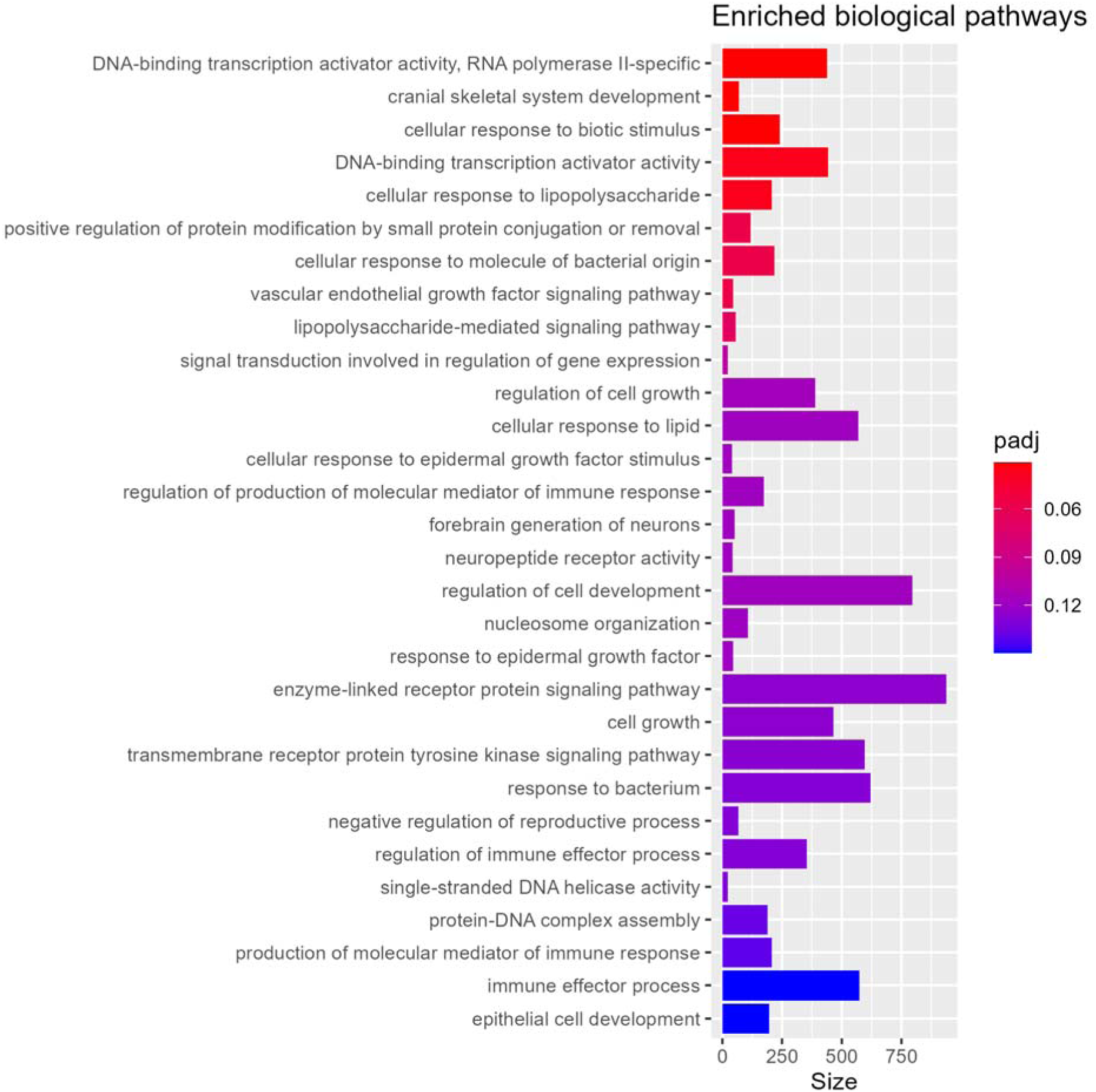
Top enriched biological process pathways in the epigenome-wide association study meta-analysis of abusive versus accidental traumatic injuries.

DNA methylation is known to play a key role in regulating promoter and enhancer cis regulatory elements (CRE).^38^ As a sensitivity analysis, we leveraged GeneHancer^32^ double elite gene-CRE associations to annotate the differential methylation observed here to promoters and enhancers with experimental evidence suggesting they affect gene expression. This confirmed that cg20544725_BC21 was located in a CRE of *AHNAK1* and *SCGB1A1*, and that cg12943082_TC21 does, in fact, affect *CCL26* expression.

To systematically identify enriched biological process pathways among genes linked to DNA methylation loci within GeneHancer CRE, we next conducted a biological enrichment analysis using this annotation. This approach identified 151 total enriched pathways (**Supplementary Table 5**), 6 of which were shared with the initial enrichment analysis: forebrain neuron differentiation, innate immune response, cellular response to nitrogen compound, growth factor binding, and response to fibroblast growth factor. The top three enriched pathways (q < 0.01) involved small GTPase–mediated signaling and kinase regulation, including guanyl-nucleotide exchange factor activity, GTPase regulation, and regulation of protein and receptor kinase activity, indicating altered control of intracellular signaling dynamics. Among the remaining pathways (q < 0.25), results indicated broad involvement of injury-response and pediatric developmental processes such as regulation of morphogenesis, ischemic response, and bone morphogenic protein signaling. Strong enrichment of neurodevelopmental pathways such as forebrain neuron differentiation, oligodendrocyte development, and synapse organization was also observed. Other prominent themes included vesicular and lysosomal–endosomal trafficking, cytoskeletal remodeling and cell motility, as well as metabolic, endocrine, mitochondrial, apoptotic, and immune regulatory processes, consistent with altered intracellular transport, structural dynamics, and stress-response mechanisms.

## DISCUSSION

In this EWAS meta-analysis of buccal cell samples prospectively collected from 175 children <4 years of age with traumatic injuries, fractures, and TBI, we identified 80 DNAm loci associated (q < 0.25) with abuse determination. Absolute average methylation rates at the top three meta-analysis loci were changed by 2.31%, 2.77%, and –13.50% among abusive compared to accidental injuries, representing fold changes of 1.39, 1.31, and 0.54, respectively, and annotated to regions of the genome associated with the expression and regulation of *AHNAK, SCGB1A1*, and an Alu element. Among other top hits at q < 0.10 *CCL26*, *LAMP1*, *RGS7* and *MIATNB* were additionally implicated. Gene set enrichment analyses at near-promoter or exonic regions identified numerous biological processes overrepresented in children with abusive compared to accidental injuries, including those pertaining to cranial skeletal system and connective tissue development, neural structure and function, immune regulation, gene expression regulation, and metabolism. Enrichment analysis of DNAm loci located in cis-regulatory elements implicated various pathways relevant to post-embryonic tissue development, neuronal and glial development, neurotrophic, metabolic, and apoptotic signaling, and immune regulation. Encompassing a diverse array of target tissues throughout the body, some specific to the traumatic injury itself and others indicative of more systemic changes to post-embryonic development, transcriptional and immune regulation, metabolism, and neurodevelopment, these responses underscore the clinical utility of buccal cells as an informative and accessible surrogate tissue for epigenome-wide association studies.^39^

Our findings are consistent with multiple prior studies revealing child maltreatment to be associated with altered patterns of DNAm that may subsequently serve as mechanisms of disease.^7^ Prior investigations into the biological effects of child maltreatment focused on non-clinical samples of older children and adults have identified and prioritized further investigation of candidate genes functionally relevant to the hypothalamic-pituitary-adrenal (HPA) axis, such as *NR3C1*, *FKBP5*, *POMC*, the *CRH* family, *SLC6A4,* and the serotonin system.^7,8,10,40,41^ We observed only nominal associations (p < 0.001-0.05) at HPA– and serotonin-related loci in a study sample of those less than four years old. The use of the GeneHancer annotation was able to identify cg00277048_BC21 in a CRE of *POMC*, and narrowly missed our q < 0.25 significance cutoff. However, it is possible that robust differential methylation at HPA-related genes is tissue-specific^42^ or occurs only at older ages than studied here in response to longer-lived maltreatment with additional neurodevelopment, a pattern observed with *OXT* DNAm.^43^

Adopting a broad approach to examine methylation across the entire genome, this EWAS identified key differences in DNAm at multiple loci that have not previously been studied as candidate genes and appear novel to the child maltreatment literature. First among these was annotated to an Alu element. Alu elements are the most abundant SINEs in humans and represent non-autonomous retrotransposons that require L1-retrotransposon machinery to function. A wealth of evidence has demonstrated that Alu elements modulate gene expression at the post-transcriptional level via multiple mechanisms, contributing significantly to the evolution of the human genome.^44^ Alu methylation is particularly dynamic during development, throughout aging, and in the setting of cancer.^45^ Typically, Alu elements are characterized by hypermethylation to repress retrotransposition, and hypomethylation of Alu elements has been associated with tumorigenesis.^46^ The significance of increased methylation at this particular Alu in our study remains unclear but may be an adaptive or stress response to ensure protein-coding transcript integrity or genomic stability. Notably, a serum-based case-control study of post-traumatic stress disorder in US service members deployed to Iraq or Afghanistan identified increased methylation at four Alu elements in cases compared to pre-deployment controls.^47^ Another study identified an association between increased oxidative stress and increased methylation of Alu elements in blood leukocytes relative to healthy controls as well as in musculoskeletal tumors relative to surrounding tissues.^48^

Our second most significant hit was annotated to intron 1 of *ENSG00000295816*, an enhancer long non-coding RNA antisense to anchor holding nucleoprotein associated with kinase (*AHNAK*), the gene coding for a large, three-domain scaffold protein expressed in a wide range of tissues including epithelial cells, muscle cells, and neurons. *AHNAK* has been implicated in many disparate processes such as cell structure and migration, cell-cell anchoring, blood-brain barrier formation, cardiac calcium channel regulation, tumor suppression, immune function, and DNA repair.^49^ This locus also serves as a CRE for *SCGB1A1*, an anti-inflammatory protein implicated in the pathophysiology of asthma.^50^ Our third key finding was annotated to intron 6 of *PTPRN2,* thought to be involved with insulin and neuron signaling and whose differential methylation in intron 2 or copy number variation has been implicated in childhood obesity^51^ or child neurodevelopmental disorders,^52^ respectively.

Another differentially methylated locus, cg16000386_BC21, mapped to a short interspersed nuclear element (SINE) repeat on the negative strand antisense to long non-coding RNA myocardial infarction-associated transcript neighbor (*MIATNB*). Though relatively less studied, *MIATNB* has been associated with the pathogenesis of dry eye disease and rheumatoid arthritis, the HIV-1 reservoir in CD4+ T-cells, susceptibility to febrile enteric bacterial infections, development of gestational diabetes, aging of alveolar macrophages, and prognosis of renal cell carcinoma.^53–59^ We also observed differential methylation at cg09283750_TC21, located with 2500bp of *FAM135B*’s transcription start site. A previous study identified an association between maternal experience of emotional abuse and offspring cord blood differential methylation at cg05486260, located in intron 9 of *FAM135B*, but we failed to replicate this finding in our study (q = 0.97).^60^ Other top hits mapped to the exon 5 of *LAMP1*, an essential component in lytic granule function for natural killer cell cytotoxicity^61^; to an enhancer within 1500bp of the transcription start site of *CCL26,* a cytokine that recruits eosinophils and basophils^62^; and lncRNA antisense to *GZMK*, an initiator of the complement cascade in inflamed tissues.^63^ Together this triad, along with enrichment of immune-related pathways, strongly suggests differential methylation plays a key role immune regulation for those with abusive injuries. Epigenetic dysregulation of *LAMP1* may have particularly deleterious consequences on neurodevelopment as elevated LAMP1 mRNA and protein blood levels have previously been associated with autism spectrum disorder in young children.^64^ Our last differentially methylated hit mapped to Intron 10 of *RGS7*, a GTPase essential for synaptic transmission by regulating GABA_B_R-GIRK signaling in hippocampal pyramidal neurons^37^ and whose knockout in mice inhibited learning and memory^65^ and modified morphine reward and worsened withdrawal.^66^ Together with the robust enrichment of GTPase-related cell-signaling, our results implicate core changes to inhibitory synaptic signaling that could broadly affect cognition, addiction susceptibility, memory, and learning.

We found multiple biological processes to be enriched among promoter-associated methylation loci for abusive as compared to accidental injuries. Several of the identified pathways involve structural and functional changes to tissues directly affected by the physical injuries studied, including cranial skeletal system development, connective tissue development, and neurogenesis pathways. By additionally leveraging the GeneHancer database to annotate DNAm loci to CREs, we sharpened our ability to identify functional epigenetic changes and tested for biological pathway enrichment among their target genes. GTPase-related cell signaling, forebrain neuron differentiation, innate immune response, and growth factor binding were the among the shared enriched pathways using both approaches. Other overrepresented pathways were relevant to long-term disease processes such as epigenetic regulation and endocrine and growth signaling that have been similarly overrepresented among individuals reporting early experiences of physical abuse. Previous studies have linked childhood maltreatment and other early adversities to chronic medical conditions spanning multiple organ systems, from mental illness to cardiovascular and autoimmune diseases, diabetes, cancer, and even premature mortality.^2,4^ Though few studies to date have examined the epigenetic mechanisms underlying this association, and rarely in young children with abusive injuries,^7,67,68^ our findings build upon earlier work to suggest a direct link between physical abuse and epigenetic responses related to central nervous system development, cytoskeletal pathways and apoptosis, cardiovascular function, immune responses and inflammatory signaling, regulation of gene expression, and cell biosynthesis and metabolism. Future large, prospective studies are needed to further characterize the role and directionality of these epigenetically-altered pathways in functional disease processes associated with early childhood abuse.

Our findings should be interpreted within the context of the study’s limitations. This meta-analysis incorporates DNAm data from three observational studies enrolling children of varying ages with different types of traumatic injury. Though not all-encompassing, this distribution reflects the typical diversity of age and injury presentations along the spectrum of physical abuse commonly encountered in clinical practice, and we observed little evidence of meta-analytic heterogeneity. However, these findings may not be generalizable to children with other forms of maltreatment. In all three studies, DNAm was measured at a single time point, raising the possibility that observed differences were driven by exposures or experiences preceding the abusive injuries themselves or as an adaptation to maltreatment. Future studies with longitudinal follow-up are needed to clarify the time course of methylation differences, including when they are first established and whether they persist with aging, development, and changes in the child’s environment. Importantly, DNA methylation is only one form of epigenetic regulation; others include histone modification, chromatin remodeling, and non-coding RNA-associated gene silencing.^69^ Though more complex and cost-prohibitive on the scale of our prior studies, experimental methods characterizing these complementary mechanisms may yield additional insights into the biological embedding of early traumatic experiences.

Overall, our results suggest that child physical abuse may not only affect proximal injury responses such as cranial skeletal system development, connective tissue development, and several neuron-related pathways, but also have lasting systemic effects on transcriptional and immune regulation, cell signaling, metabolism, and neurodevelopment. These findings underscore a critical need to improve our understanding of the biological pathways that translate early adversity into long-term health consequences. Further investigation of the role of epigenetic modifications and interactive bioregulatory pathways that shape lifelong trajectories of health and disease can ameliorate pediatric morbidity and mortality from child maltreatment and improve health outcomes among young children with abusive as well as accidental injuries.

## Declarations

### Acronyms

DNA: deoxyribonucleic acid

TBI: traumatic brain injury

TSS200: within 200 base pairs of a transcription start site

## Data availability statement

The datasets generated and/or analyzed during the current study are available from the corresponding author on reasonable request. Code to produce the analyses in this manuscript are available through GitHub (https://github.com/…).

## Acknowledgements

We thank the participants who provided biospecimens for this study. This research was supported by the Stanley Manne Research Center of Ann & Robert H. Lurie Children’s Hospital of Chicago Visionary Award and Fellow Research Scholar Award, Ann & Robert H. Lurie Children’s Hospital of Chicago Foundation Innovator Award, Ann & Robert H. Lurie Children’s Hospital of Chicago Pediatric Physician Scientist Research Award, Grainger Research Initiative Fund in Emergency Medicine at Ann & Robert H. Lurie Children’s Hospital of Chicago, and the HERCULES Center (NIEHS P30ES019776).

## Author contributions statement

**Kyle Campbell:** Conceptualization, Methodology, Software, Validation, Formal analysis, Investigation, Data Curation, Writing – Original Draft, Visualization

**Audrey Raut:** Conceptualization, Methodology, Data Collection, Sample Collection, Investigation, Funding Acquisition, Writing – Original Draft, Project Management

**Kelsey Julian:** Project Management, Writing – Review and Editing

**Kim Kaczor:** Conceptualization, Methodology, Funding Acquisition, Writing – Review and Editing

**Kathi Makoroff:** Conceptualization, Methodology, Writing – Review and Editing

**Todd M. Everson:** Conceptualization, Methodology, Software, Validation, Formal analysis, Investigation, Funding Acquisition, Data Curation, Writing – Review and Editing, Resources, Supervision

**Mary Clyde Pierce:** Conceptualization, Methodology, Data Collection, Sample Collection, Investigation, Funding Acquisition, Writing – Review and Editing, Supervision

## Supplementary Figures and Table

**Supplementary Figure 1.**
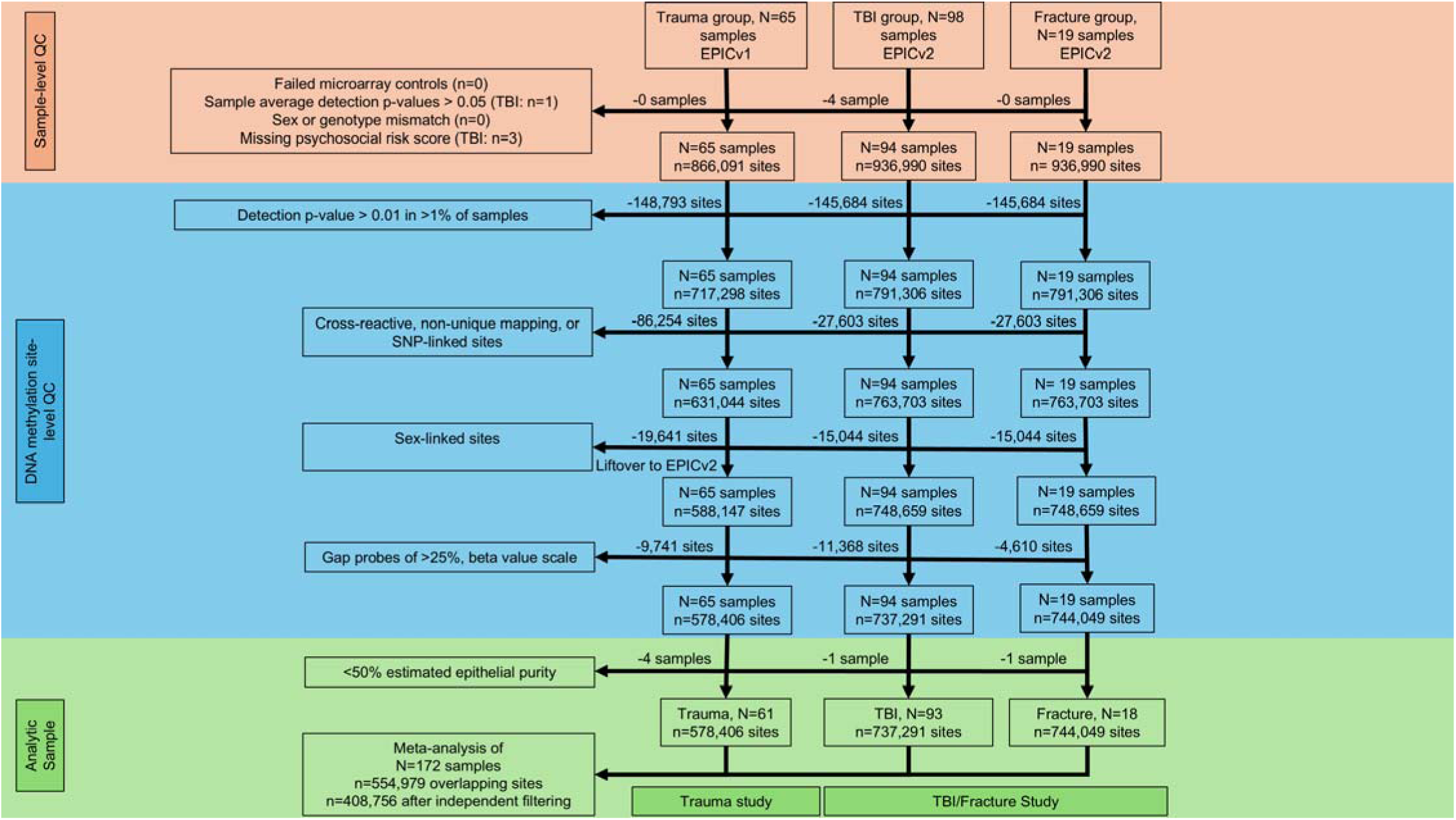
Overview of quality control procedures presented as a flow chart for each study sample. Sample-level quality control exclusions painted in orange. DNA methylation site-specific exclusions painted in blue. Final analytic sample after exclusion of low predicted epithelial cell origin painted in blue.

